# “If you’ve ever watched Pearl Harbour and that nurse comes out and everything is spinning, and you just don’t know where to go”: A qualitative study on the unseen struggles and resilience of LTCF staff in England during the COVID-19 pandemic

**DOI:** 10.1101/2025.05.23.25328140

**Authors:** Natalie Cotterell, Danni Collingridge Moore

## Abstract

**Background:** The COVID-19 pandemic adversely impacted health professionals’ mental health and emotional wellbeing, however less research has examined the impact of the pandemic on LTCF staff. There is limited research on the emotional wellbeing of adult social care staff, particularly within the context of the government guidance and restrictions. This study aims to explore the impact of the COVID-19 pandemic on the mental health and emotional wellbeing of LTCF staff in England.

**Methods:** Semi-structured, qualitative interviews with 24 staff members working in eight LTCFs based in the north-west of England. Data was collected initially to explore experiences of staff members working in LTCFs during the COVID-19 pandemic, and a secondary analysis of the data focused on mental health and emotional wellbeing was conducted using thematic analysis.

**Findings:** Five central themes were conceptualised: burnout and trauma (including initial impact on mental health and long-lasting effects on mental health); efforts to cope (passive coping strategies and active coping strategies); positive impacts on staff wellbeing (including building resilience and preserving the routine of daily life); impact on personal/home life (including protecting ones family, changing family relationships, and less work/life balance); and availability of support (including informal support and support from external services).

**Conclusions:** The COVID-19 pandemic had a predominantly negative impact on LTCF staff’s emotional wellbeing, partly due to a lack of external support and appreciation. This impact continues to affect many LTCF staff who have not been able to access useful or appropriate support. This research highlights the importance of developing organisational resilience within such facilities and supporting LTCF staff to emotionally cope with the challenges of future pandemics.

## Background

The COVID-19 pandemic brought numerous challenges for long-term care facilities (LTCFs) across the globe (1). In the UK, the ongoing COVID-19 inquiry is reviewing how government policies designed to protect adult social care impacted LTCFs, particularly in terms of outbreaks, COVID-19 related deaths and distress for residents, relatives and staff members (2). In total, there were a recorded 39,265 COVID-19 related deaths across LTCFs in England between April 2020 and March 2021 (3). By May 2020, LTCF residents accounted for 54% of all COVID-19 related deaths in England (4). Previous research has demonstrated that the increased mortality among LTCF residents was a source of distress for care workers (5). In addition, care workers were managing multiple demands relating to the COVID-19 restrictions on visitation, social distancing and infection prevention control, staffing shortages, vaccinations, continuing routine care, and managing patients discharged from hospital (6). International studies have further reported care staff experiencing higher incidences of challenging behaviour amongst residents, reductions in support from families and outside agencies and increased work and emotional loads (7–9).

Despite the known challenges, the effects of the pandemic on the emotional wellbeing of staff working in LTCFs have received less academic attention than the impact on frontline medical staff (10, 11). A growing body of research has demonstrated the negative impacts of the pandemic on adult social care staff. A European survey conducted in the UK, Sweden, Italy, and Germany found that stress and anxiety levels were highest amongst care staff in the UK (12). The authors state that this was partially explained by the lack of organisational support and staff voice compared to other countries. More recently, a systematic review, examining 14 studies related to exploring experiences of LTCF staff during the COVID-19 pandemic, highlighted that staff experienced a number of mental health concerns (13). With five of the studies coming from the UK, it was found staff experienced heightened anxiety, depression, grief, and exhaustion; however, all but one of the UK studies included only LTCF managers (5, 14–17). The study that included a range of care staff reported on the triggers of mental health problems amongst frontline LTCF and domiciliary workers, which included: fear of infection; lack of recognition; lack of guidance; unsafe hospital discharge; death and loss of professionals; unreliable testing and delayed results; and shortage of staff (5).

In the UK, the extent to which formal support offered to LTCF staff throughout the pandemic by the government was effective is debatable (12). The support offered included a mobile application called ‘*The Care Workforce App*’, launched in May 2020, and provided various toolkits and resources to support mental health and wellbeing (18). However, to date there has been no evaluation of the app, making it unclear to what extent the app supported care workers with their emotional wellbeing.

Some researchers have suggested that the concept of resilience may have mediated the impact of the pandemic on emotional wellbeing amongst care workers (19). Resilience can be defined as “*the process of effectively negotiating, adapting to, or managing significant sources of stress or trauma. Assets and resources within the individual, their life and environment facilitate this capacity for adaptation and ‘bouncing back’ in the face of adversity*.” (20). Previous research has demonstrated that personal resilience can prevent burnout among clinicians working in wider healthcare settings (21). During the COVID-19 pandemic, a moderate level of resilience has been reported in a survey of LTCF nurses, with key predictors of increased resilience being older age and higher levels of education (22). Furthermore, in research focused upon the role of resilience among UK care staff during the pandemic, qualitative interviews identified staff’s adaptive and proactive responses to the impact of overwhelming workloads, anxiety, and distress (23).

The aim of this paper is to expand further on the current research base, capturing the perspective of adult social care staff based in LTCFs, exploring their experiences of working through the COVID-19 pandemic and its impact on their emotional wellbeing and mental health.

## Methods

### Study setting

The data analysed within this paper were originally collected in a mixed-methods case study that examined the implementation of government issued COVID-19 policies within LTCFs in England. The data were collected during 24 interviews with staff from eight LTCFs based in six local authorities across north-west England. The Enabling Research in Care Homes Network distributed information about the study to facilities which had expressed an interest in taking part in research studies (24). Inclusion criteria for participation in the study included registration with the Care Quality Commission (CQC) to provide residential care to older adults aged 65 and over and were active during the COVID-19 pandemic. Facilities that expressed an interest in taking part in the research were contacted by the research team, and staff members who were interested in taking part in an interview were provided with an information pack detailing what participation would involve. Inclusion criteria for staff participation included employment in the facility during the pandemic, aged 18 years and over and able to conduct the interview in English.

### Data collection

In each LTCF, between two and four staff members were recruited to take part in semi-structured qualitative interviews. Information packs handed out by the LTCF manager to staff members who met the prespecified inclusion criteria, and purposefully approached staff members from a variety of roles at the request of the research team. All interviewees provided written and verbal informed consent to take part. A broad interview topic guide was developed, and no specific questions asked about the emotional wellbeing of staff, although this topic emerged as significant in every interview. All interviewees received a £25 gift voucher following their interview to thank them for taking part, as recommended by previous research studies conducted in adult social care settings (25). In total, 24 LTCF staff members who represented a range of different roles within each setting, including managers, care assistants, housekeepers, and activity co-ordinators, were interviewed. Full participant characteristics are presented in Table 1.

**Table 1:** Demographic and occupational characteristics of interview participants.

| **Variables** | **Mean** | **(range)** | **N** | **%** |
| --- | --- | --- | --- | --- |
| Age | 45.9 | 28-68 |  |  |
| Gender - female |  |  | 21 | 87.5 |
| Ethnicity - white British |  |  | 21 | 87.5 |
| English first language |  |  | 22 | 91.7 |
| Years working in direct resident care (1) | 21.3 | 5-46 |  |  |
| Years working in LTCFs | 18.2 | 5-46 |  |  |
| Full-time staff |  |  | 19 | 79.2 |
| **Current position or role** |  |  |  |  |
| Registered LTCF manager |  |  | 5 | 20.8 |
| Deputy LTCF manager |  |  | 2 | 8.3 |
| Head of operations |  |  | 1 | 4.2 |
| Clinical manager |  |  | 3 | 12.5 |
| Senior care assistants / CHAPs team leader (2) |  |  | 6 | 25.0 |
| Care assistant |  |  | 3 | 12.5 |
| Activity co-ordinator |  |  | 1 | 4.2 |
| Housekeepers |  |  | 3 | 12.5 |
| Residents being provided direct care | 39.9 | 12-118 |  |  |
| **Highest level of education you have completed** |  |  |  |  |
| Tertiary level (higher vocational training or university) |  |  | 10 | 41.7 |
| Higher-secondary level (up to A level or equivalent) |  |  | 6 | 25.0 |
| Lower-secondary (up to GCSE or equivalent) |  |  | 8 | 33.3 |

(1) The mean number of residents each interview participant provided care to on a daily basis. (2) CHAPs - Care Home Assistant Practitioners.

Semi-structured interviews were conducted with staff members from each facility between October 2023 and March 2024. The interviews were primarily conducted by NC, ranged from 40 to 60Lminutes in length and were either conducted online using Microsoft Teams or in person at the LTCF depending on the preferences of the participant. During the interview, participants were asked to reflect on their role at the facility during the pandemic, how their role changed in relation to the implementation of government issued policy recommendations and any wider reflections they had on working during the pandemic.

### Data management and analysis

The interviews were recorded using Microsoft Teams and transcribed using Microsoft Word. The transcripts were anonymised, and each participant was given unique identification codes. Each interview transcript was analysed using thematic analysis following the five step approach developed by Braun and Clarke: a) familiarisation with the data, b) generating initial codes, c) searching for themes, d) reviewing themes and e) defining and naming the themes (26). ATLAS.ti was used to store, manage, and organise interview transcripts, and the COREQ checklist was used to guide the reporting of the research (27).

### Interviewee characteristics

In total, 24 LTCF staff members who represented a range of different roles within each setting, including managers, care assistants, housekeepers, and activity co-ordinators, were interviewed. Full participant characteristics are presented in Table 1.

### Ethical considerations and researcher reflexivity

Ethical approval was obtained from the Lancaster University Faculty of Health and Medicine Research Ethics Committee (reference: FHM-2023-3368-RECR-3) in July 2023. Written and verbal informed consent was taken from LTCFs and staff members; both held the right to withdraw their consent to take part and any data provided at any point prior to data analysis. A distress protocol was in place during the interviews in the event of the interviewee feeling upset or distressed while discussing their experiences. Each interviewee received a signposting and debriefing information sheet following the interview. Facility identifier codes have been removed from quotations to ensure individuals cannot be identified from the information provided. The authors identify as female researchers and were employed solely on the study; both hold post doctorate degrees and undertook additional training on conducting qualitative research, including conducting online interviews and analysing qualitative data.

### Findings

The findings highlight the extent to which the COVID-19 pandemic had a negative impact on both the short term and long-term emotional wellbeing of LTCF staff, contributing to a deterioration of their mental health. However, the findings also demonstrate an undercurrent of mental resilience amongst some staff. Five central themes were conceptualised: burnout and trauma; efforts to cope; factors facilitating self-empowerment; impact on personal/home life; and availability of support. The sub-themes are presented in Table 2.

**Table 2:** Overview of core themes including description of theme and sub-themes.

| Core theme | Sub-theme(s) |
| --- | --- |
| Burnout and trauma | Initial impact on mental health<br>Long-lasting effect on mental health |
| Efforts to cope | Passive coping strategies<br>• *Avoidance coping*<br>• *Distraction coping*<br>Active coping strategies<br>• *Reframing*<br>• *Social coping* |
| Factors facilitating self-empowerment | Building resilience as a carer<br>The power of routine in difficult times |
| Impact on home life | Protecting one's family<br>Changing family relationships<br>Less work/life balance |
| Availability of support | Informal support<br>Support from external services |

### Burnout and trauma

#### Initial impact on mental health

Initial emotions when the first COVID-19 lockdown was announced in the UK in March 2020 ranged from fear and panic to relief amongst interviewees. The government announcement of lockdown shocked some interviewees, as they felt it was unexpected and sudden – particularly among those who believed it was a ‘Chinese problem’ that would not affect the UK. In contrast, others had already been involved in preparations to protect residents from the virus; for example, some LTCFs had chosen to close the facility to visitors prior to the announcement. These interviewees reported a sense of relief as the government took action, meaning the decision to restrict visitation was then outside of their control:

> *“When the lockdown happened, there was a bit of relief that somebody else has made that decision, that it’s no longer us trying to enforce it, because it did feel a bit like some families were really happy and grateful with residents [not being visited]. Others were very upset and annoyed that we’d put these restrictions in place, really. So when the government did it, it wasn’t up to us anymore*.*”*
>
> P017

The lockdown, alongside government restrictions, meant that staff were dealing with overwhelming and growing workloads and responsibilities. Interviewees reported experiences of burnout, detailing the toll it took on their mental health at the time. Participant 021 explained how it was the hardest time of their life, affecting her both mentally and physically:

> *“I know for myself, really, it’s the hardest time of my life that I’ve ever had to deal with. And I say it’s the responsibility, I remember doing a FaceTime with my niece. She said you look dreadful. You look awful. You look absolutely*… *and I said yeah I feel it. I just feel, you know, such a responsibility really to these people and the families and you know, we just get every so often this guidance that contradicts itself or was too late and it was difficult*.*”*
>
> P021

Participant 021 described how the regularly changing guidance that they deemed to be often contradictory contributed to the stress experienced. Participant 023 further echoed the point of feeling burnout and overwhelmed:

> *“Yeah, we had to because we got ourselves drained that much. It was quite shocking actually. I think that’s why some of us became poorly as well, yeah. We’re bouncing off each other to work all the hours we could without having to get agency (staff) all the time, you know, instead of letting it spread. It was hard*.*”*
>
> P023

In particular, managers of LTCFs reported having increased and overwhelming responsibilities that contributed to deteriorating physical and mental health. Participant 021 explained how ‘terrifying’ they found managing the LTCF during the pandemic as they felt like they was responsible for residents’ deaths and would be blamed for them dying with no family visitation:

> *“So for me personally it was terrifying. It was a huge responsibility. It felt like as a care home, we were completely on our own. Nobody was supporting us or telling us what we should and shouldn’t do. […] to me that felt like I could kill people’s family. And on top of that, the fact that their families weren’t visiting. And I had that responsibility. I felt terrible*.*”*
>
> P021

They later commented on how this responsibility interrupted their sleep and limited their self-care which had a consequent effect on their mental health. Further, they reported that feeling like the LTCF was ‘completely on our own’ was an additional burden and contributed to how overwhelming the responsibilities she had felt. Multiple responsibilities contributed to the exhaustion reported by all LTCF managers in particular. Another LTCF manager described how they had made a bed for themselves at the LTCF as they were working ‘day and night’ due to staff shortages; and discussed how they felt responsible for minimising the spread of the virus to both residents and their own elderly parents. They reported going into work to do paperwork even when they felt extremely unwell as they tested negative for COVID-19, though later found out through an antibody test that they had had COVID-19:

> *“All my nurses got it at the same time. I was the only one left, working day and night and I felt ill myself, I got tested but I was negative which later proved that I actually was positive. I just didn’t show positive results on the LTCF I only found out through a blood test that I did actually have it cause the illness was so horrible I thought, gosh, I mean, I was coming to work, was on exhaustion. I had a bed here and I was exhausted and I remember coming to work one day. Just crazy, you know? And I’m coming to my death here, you know?*.*[…] I still did it [paperwork]. I still worked and did it, you know? But yeah, so but…I was shattered, absolutely shattered*.*”*
>
> P006

#### Long-lasting impact on mental health

For some interviewees, the impact of the pandemic on their mental health was longer-lasting than during the pandemic alone. Several interviewees reported that they continue to struggle to process exactly what happened to them during the pandemic and its effect on their mental wellbeing. One interviewee reported that they regularly still have reoccurring dreams involving memories of the residents who passed away; another reported having no memory of the first lockdown. Participant 022 admitted that they had not thought about the period around the pandemic since it happened, and the interview was one of the first times they had spoken about it. Taking part in the interview made them feel emotional, which took them by surprise:

> *“I think it did have an impact on people’s lives and it’s only when you really talk about it that you realise, you know, but yeah, it did affect me and I’m becoming quite emotional about it, which is a bit strange, but we’ve got to take positivity away from it, haven’t we? You know as well. Yeah, that’s a hard one*.*”*
>
> P022

In contrast, there was sometimes a sense that a sort of positivity had to be gained from the pandemic, particularly amongst interviewees who were ‘getting on with it’ or who possibly had not fully processed the experience. Participant 012 struggled with remembering the details of what happened during the pandemic and admitted they had ‘blocked it out’; however, they spoke about how they felt the ‘damage had already been done’ and no retribution or lessons learned by the government could reverse that:

> *“I’ll be honest, I can’t remember back then. I think it just all happened. It’s only now that things have calmed down that what they talk about, what things could have been done better to protect LTCFs, the elderly and vulnerable. So obviously there’s now a COVID inquiry going on in there, but I don’t really pay much attention to that now because the damage was done back then*.*”*
>
> P012

### Efforts to cope

#### Passive coping strategies

Many interviewees implemented passive coping strategies to deal with the impact of the pandemic on their mental wellbeing, helping them to temporarily evade the associated potentially negative emotions and preventing them from directly confronting the challenges encountered. Two types of passive coping strategies were employed by interviewees: avoidance and distraction.

*Avoidance coping:* Interviewees who used avoidance coping tended to withdraw socially and emotionally from loved ones, attempting to avoid both experiencing and sharing anxiety and fear.

This appeared to be particularly common amongst managers, who spoke about wanting to avoid burdening staff with their own fears so kept them to themselves. However, care staff also spoke about how they did not want to burden their family and friends and did not share their feelings or experiences with them. Participant 013 talked about avoidance coping, reporting how they did not cry in front of their partner and did not speak about work at home:

> *“I remember going home every night, getting in the bath and just crying. I’m looking in the mirror and just crying. I wouldn’t cry in front of my partner because I didn’t want him to see how it how it made me feel so you try and stay strong at work, and then you try to stay strong at home, but somewhere that has to come out*.*”*
>
> P013

Distraction coping: Distraction coping is considered to be more active than avoidance as individuals are making an attempt to manage the stress (28); however, individuals were ‘keeping busy’ rather than confronting any negative feelings, suggesting there was still some level of avoidance in confronting and processing emotions. Participant 012 spoke about how they felt a strong duty of care towards both residents and colleagues and therefore was too busy to think about their mental wellbeing; thus, their work became a distraction:

> *“It was hard but we had to do what we had to do, because of our, you know, we had to be there for the residents and do what we could. […] I was kept busy, so didn’t have time to think about it all, or how it affects me*.*”*
>
> P012

#### Active coping strategies

Some interviewees employed more active coping strategies, attempting to alleviate potentially negative emotions. Most employed were reframing and social coping – both of which will now be explained, in turn.

##### Reframing

One interviewee acknowledged the potential negativity of the pandemic including the restrictions and they then challenged this thought, changing the way they considered the impact of the pandemic:

> *“All I thought was just get on with this. There is nothing to complain because the world is stuck. There is nothing, nothing going on around the world. Everything was stuck. A lot of people are losing their beloved ones*.*”*
>
> P009

By referring to people losing their loved ones, the interviewee appears to be thinking about how their situation could have been worse. They mentioned that they were not able to go out and do the things they wanted to do after work during the restrictions, yet reframed this by acknowledging how the whole world was ‘stuck’ and so they were not missing out as such.

##### Social coping

Several interviewees discussed how socialising with colleagues aided their coping during the pandemic by boosting their mental wellbeing through bonding via a shared connection. Participant 001 spoke about how they became close with colleagues during the pandemic as they were all going through a shared experience:

> *“Me and my four, five of us that became like close best friends, right. We’re just doing quiz nights on teams. […] It wasn’t necessarily talking about it as such it was just all four of us were going through the same situation at work. So we can all kind of relate if needs be*.*”*
>
> P001

They reported how this close connection helped support their mental wellbeing, despite not explicitly discussing their anxieties with their colleagues.

### Factors facilitating self-empowerment

#### Building resilience as a carer

Some staff reported that they had become more resilient, overcoming personal challenges during the pandemic which may have protected them from deteriorating mental health. Many interviewees spoke about how they felt their experience had prepared them mentally for any future pandemics, as well as helping them to cope while working during the COVID-19 pandemic. Participant 007 spoke about how they felt ‘used to death’ because they had witnessed so many resident deaths during the pandemic; they explained how dealing with death does not bother them as much as it did pre-pandemic:

> *“I don’t want to sound like heartless, but now when someone dies it doesn’t affect me as much, you know. I don’t mind going in and washing them up, you know, because I’ve seen so much of it. But then there’s, like, young girls whose just started here and weren’t here for COVID and they won’t even go in a room when someone died and that, you know, I think I had to deal with that when I was your age*.*”*
>
> P007

LTCF managers mainly held the attitude of ‘just get on with it’ due to their strong sense of duty of care for both residents and staff and saw managing through a pandemic as part of their job role. Participant 008, an LTCF manager, remembered that they almost ‘gave up’ and left the profession, though a ‘pep talk’ from a CQC inspector helped them to feel more confident and able to deal with the challenges:

> *“So it was a tough time. I nearly gave up. I remember speaking to the inspector and I said, oh, I think I’ll have to resign. It’s too hard being the manager and being a nurse. And I said doing either maybe. And he said, oh, don’t do that, you know, you can do it. And yeah, gave me a pep talk and it worked. I’m still here*.*”*
>
> P022

Other managers felt they had to be ‘the strong leader’ despite sometimes wanting to give up and get an easier vocation. This may have helped them cope with the challenges of the pandemic, contributing to their personal and organisational resilience. Participant 006, reported putting on a brave face, role modelling calmness and optimism so that staff felt confident in the management:

> *“You’ve got to come out of this door, and you’ve got to be… if you’re down, the staff are down. If you’re frightened, the staff are frightened. You’re their home, their mother hen really. They look up to you. They rely solely on you, and it’s a big responsibility, you know*.*”*
>
> P006

Most LTCF managers reported that they would feel more confident dealing with a future pandemic due to their experience of managing a facility through the COVID-19 pandemic; though all managers hoped that there would not be another pandemic in their lifetime. The manager of LTCF 004, however, admitted that they would likely leave their job if a future pandemic occurred due to the immense responsibility they had felt:

> *“I personally wouldn’t want to go through it. So, I think I’d leave. I just wouldn’t want that responsibility again. It was so horrendous*.*”*
>
> P021

Thus, they did not want to manage another pandemic and later questioned whether they could ‘cope’ with another pandemic, bringing their personal resilience into question.

#### The power of routine in difficult times

Many interviewees spoke about how work preserved the routine of their daily life, having a positive impact on their wellbeing by maintaining a level of ‘normality’ and providing opportunities for solace. Staff reported feeling fortunate that they had somewhere to go and something to do during the lockdowns, compared to those who were furloughed or worked from home full-time. Some interviewees reported that reduced traffic made their commute more enjoyable as it was quicker to get to work and more tranquil to experience. Participant 001 recalled how continuing to work throughout the pandemic maintained their mental wellbeing:

> *“Think I was doing like six nights a week or something just to*… *I couldn’t do anything, couldn’t be with my family, couldn’t go out anywhere…so work was keeping me sane*.*”*
>
> P001

It is possible that being in a caring role and having to travel to work every day contributed positively to staff’s wellbeing by helping them to maintain a sense of normality. Participant 019 echoed this point and explained how they felt their job brought some normality to their life during lockdowns compared to those working at home:

> *“It brought some normality to your life because actually a lot of people were furloughed as well at the time. Our place was just shut down. Yeah, and I think it was probably worse for them being locked down. But we had some, even though it was abnormal, we had a bit of normality to our lives through working through the lockdowns and so in some ways that was better. Because I think a lot of people’s mental health suffered when they were not working, people, even people who were working from home, yeah. I know there was an impact on those. So in some way, the other benefit was that normality in your life*.*”*
>
> P019

### Impact on home life

#### Protecting one’s family

In addition to having a duty of care for residents and colleagues, care staff also spoke about how the COVID-19 pandemic impacted their personal lives and the associated emotions. Interviewees mentioned how they separated work and home as much as possible to keep both residents and their loved ones safe. They made personal sacrifices such as not visiting their elderly parents even when some of the restrictions had eased. Participant 014 remembered how ‘strange’ they felt it was to not give their children a hug when they returned from work, instead they went immediately upstairs and showered:

> *“When I got in [from work], before my kids could come near me, I went straight up the stairs to shower before giving them a hug. […] I’ll never forget that and what a strange time it was, I wanted to protect my kids*.*”*
>
> P014

Participant 008 echoed this point, describing how they cannot imagine telling their children that they were not allowed to give them a hug in order to protect them from the virus:

> *“I can’t imagine now telling my children when they’re my age that we were. I weren’t allowed to hug you when I came home”*
>
> P008

#### Changing relationships

Some care staff reported the COVID-19 pandemic had impacted their relationships with family and friends: sometimes positively, for others negatively. Participant 007 mentioned that their partner came to appreciate how difficult their job as a care assistant is. Prior to the pandemic, their partner did not appreciate how mentally and physically taxing their job was:

> *“We get some people who think its easy and they leave after two weeks and its not as easy as they make out. My partner used to say you just go to work and make brews all day and he doesn’t say that anymore! He knows how hard it is now*.*”*
>
> P007

Participant 007 thought this had improved their relationship by increasing their partner’s understanding of their job role. On the other hand, participant 013 found that their partner had a lack of understanding for their job and associated responsibilities during the pandemic; this caused increased arguments and stress at home, contributing to a decline in their mental health:

> *“I think that’s where I found it hard because our partners didn’t understand. So they’re going ‘well why do you have to go in?’ Well, because we have to and they just didn’t understand why you are going from 9-5 to like 8am-8pm or 8am-11pm and they just didn’t understand. So you’re getting arguments at home. ‘Why are you not home? Why this…’ and all you can say is it’s my job*.*”*
>
> P013

Participant 013 felt a strong sense of duty and so worked whenever the LTCF needed despite this placing a strain on their relationship.

#### Less work/life balance

All staff reported having less of a balance between work and home life during the pandemic, with many reporting that they regularly worked between 12 and 16 hours per day alongside picking up extra shifts. Several interviewees reported that they slept over at the LTCF when staff numbers were particularly low – this was particularly common amongst care managers or those in a senior role. Participant 009, a Unit Manager, explained how they stayed over at the LTCF for one week even on their birthday despite their friends asking them to return home:

> *“My friends were calling me saying that. Come on, come back. It was my birthday as well. I didn’t go. Yeah, it was XXth of March is when the lockdown started. I was stuck here from XXth. So next, next day is my birthday. My friends were calling, come, come back you don’t have to stay there, we want you to come back and I said there is no one. There is no one. and I can’t come just like that so I took a chance. It wasn’t exhausting. It was more like I didn’t feel it is anything physically or mentally for myself, I felt it’s … this place has to keep going. There is something going wrong, something I need to help*.*”*
>
> P009

Participant 009 felt such a strong duty of care that they felt unable to leave the LTCF with low staff numbers. They further explained how this imbalance between work and life had continued beyond the pandemic, changing their everyday routines, motivations, and interactions; consequently, impacting their emotional wellbeing:

> *“But after and during COVID, we were like spending a lot of time home. I was working every day of the week, I’d never stopped working. But whenever I get day off, me and friends we just ohh can we cook something and drink. Thats what we started doing during the COVID, so after that the pattern is still there. So it’s getting a bad habit as well, habit into the lifestyle. So it’s more like our preferences has changed, so now I’m struggling to change that. […] I have no motivation to go out, because we were stuck in the house or just work, house, work, house. That has been completely it for two years. […] To get out and about now it’s it needed a lot of motivation*…*it’s not great on my mental health*.*”*
>
> P009

Participant 009 talked of how their motivation has been lost since the pandemic over two years ago and how their habit of just leaving their home for work has become more of a lifestyle change. They reported that it had negatively impacted their mental health and increased health-risk behaviours such as increased alcohol consumption and overworking.

### Availability of support

#### Informal support

Most interviewees received informal support from their peers and colleagues, which, in some cases, was the only support available. Peer support was deemed to be sufficient by some interviewees, but not enough by others. Some LTCFs trained staff as mental health first aiders who were then responsible for providing mental health support to colleagues during the pandemic. Participant 018 recalled how the informal group they set up during the pandemic helped staff not only emotionally but also financially given that some staff were experiencing a reduced income due to the restrictions imposed on the hospitality sector during the pandemic:

> *“I’m a mental health first aider and I’ve really introduced like a well-being resilience group and trying to kind of get people because people were coming to us and like saying such and such body is like, keep picking up loads of shifts. They don’t want to be at home on their own. And I just feel like they’re really tired and such and such bodies come and they can’t afford this because they used to do pub work as well and… were like. buying a pair of shoes from a member of staff because they literally like had debts and didn’t have any other way […] a woman that works here, who whose husband worked in hospitality and they relied on tips for Christmas presents for the kids, it’s like some really heartbreaking stuff, you know? And all that would, we would probably not have ever got to that information cause people come to work, do the job, go home, you get some people that come and need support and they come and share, but we found that more people were really open to sharing. It was as if, like, we were like family really*.*”*
>
> P018

Participant 018 also described how the staff became ‘like family’ referring to the strengthening of the emotional bonds amongst the staff team due to sharing their emotional and financial struggles with one another. Many of the interviewees mentioned that their only source of support was their manager. Participant 008 described how the manager supported staff emotionally each day, emphasising the ‘togetherness’ of the team while remaining strong as a leader:

> *“We just got to get [our manager]. Yeah, she’d come in every morning and she was like, like, it’s funny now what she give us this little pet talk in the morning. And it’s like, right we’re gonna be all right. And you know. We’re gonna do this together. We’re gonna get through it. And because she must have seen our faces. It’s like we were all like wide eyed, it’s like handover, scared to death but she was scared, weren’t she?”*
>
> P008

Participant 008 also mentioned how the care providers helped out in the LTCF and got to know the staff, perhaps in an attempt to boost morale. This was generally appreciated and helped contribute to the sense of ‘togetherness’. In comparison, Participant 009 mentioned that although the managers tried to provide emotional support, they were not receiving any support themselves; the participant showed empathy for their situation:

> *“Our managers were like trying to support us, but still they didn’t have any support. They don’t have any support, so it’s more like we don’t have support, they don’t have support, no one around us were having support. It’s just get on with it*.*”*
>
> P009

#### External support

Almost all LTCFs reported a lack of practical and emotional formal, external support and only two LTCFs received external support for staff experiencing mental health challenges during the pandemic. For those who did receive external emotional support, counselling was provided by the local authority, however this was largely considered to be inappropriate due to poor timing and delivery. One registered manager said it was ‘hard to engage with’ and explained how the timing of the offer felt wrong as they had not had any time to process the trauma that they were still experiencing as the pandemic was ongoing:

> *“They [local authority] offered us some…they offered me some kind of managers counselling, and I went, but it weren’t like counselling. it was a bit bizarre because it was going on while the outbreak was still happening. And you know when you’ve been through a trauma, you’re supposed to process it and deal with it after. Well, this was this was very much whilst I was in the middle of it and I just didn’t find it very helpful at all because it weren’t*… *It weren’t one-on-one. There was a group of us. […] then you’ve got some managers who were there and they were going on about nonsense things. They hadn’t even had anyone die in their home then they were like, comparing their so-called situation to mine. And I found it. I found it really hard to engage with it. It were weird. Anyhow, I did about one or two sessions and I just didn’t bother*.*”*
>
> P005

Interviewees also mentioned that they felt there was a general lack of appreciation and support from the government, authorities, and communities, contributing to feelings of worthlessness and futility amongst some interviewees. Numerous comparisons between the social care sector and the National Health Service (NHS) were made, with care staff feeling that they were ‘forgotten about’. Participant 007 remembered a small gesture of appreciation they received from two individuals who clapped for the staff in their car park, reporting that it made them feel emotional:

> *“We were forgotten about because we weren’t NHS. I remember the clap for NHS, they used to do it, there used to be this guy and his wife and every night he used to stand on the car park and clap for us. It used to make me well up every time. We were just forgotten about*.*”*
>
> P007

Most interviewees explained how they felt alone during the pandemic with extremely limited emotional support, invoking feels of anger towards the government.

> *“I’m very angry. Very, very, very angry. I still feel angry now, at what they [government] did*.*”*
>
> P005

## Discussion

This study aimed to explore the impact of the COVID-19 pandemic on UK care worker’s emotional wellbeing. The findings highlight challenges that focus on burnout and trauma due to overwhelming workloads and lack of resources, emphasising the long-lasting effects on some interviewee’s wellbeing. Efforts to cope with the challenges of the pandemic included passive coping strategies, where some interviewees reported that they avoided showing emotions they felt or distracted themselves; and active coping strategies with some interviewees reframing their perceptions and seeking out social support. During the pandemic, staff built resilience with most interviewees claiming that they would mentally cope well in the event of a future pandemic. Some staff also mentioned that their jobs meant that they were able to preserve the routine of daily life, consequently protecting them from deteriorating mental health by maintaining a ‘sense of normality’. Staff further reported that the pandemic impacted their personal lives in various ways; interviewees living with children and/or older people reported taking further precautions to protect those who they considered more vulnerable to the virus in addition to protecting the residents of the LTCF. Some interviewees reported that their relationships with family had changed, some felt more appreciated whereas others experienced increased conflict due to the pressures of their job. Most interviewees reported no work/life balance, with some staff sleeping in the LTCFs and working 16 hour shifts; this had a consequent negative effect on mental health. Finally, despite the clear impact on staff’s emotional health there was a reliance on peer support throughout the pandemic. Peer and managerial support were sufficient for some interviewees, whereas others reported feeling that they need more emotional support. There was a consistent lack of external support from wider services and those who did receive external counselling reported that it did not improve their emotional wellbeing. Staff generally felt underappreciated and unrecognised, with feelings of anger towards the government and a loss of trust in their ability to guide and support LTCFs.

Consistent with previous research, the findings highlight the negative impact on care staff’s emotional health during the COVID-19 pandemic across LTCFs in the north-west of England. This paper adds to knowledge on how, as the crisis of the pandemic has subsided, care staff have been impacted in the longer-term by the COVID-19 pandemic. It shines a light on issues within the care sector that existed prior to the pandemic, highlighting the importance of finding feasible solutions in addition to future pandemic planning.

### Implications and recommendations

The findings within this paper have several implications: the adverse impact of the pandemic on staff’s emotional wellbeing must be prioritised, especially in the event of a future pandemic. In particular, external emotional support from wider services should be made available to all care staff to improve staff’s mental wellbeing. This support must be timely, appropriate, and effective. It is recommended that future researchers engage with LTCF staff to co-design interventions designed to support emotional health alongside mental health experts. Furthermore, resilience has been demonstrated to protect LTCF staff against deteriorating mental health (19); thus, it is important to ensure all care staff are able to further build and develop personal resilience. For example, this may involve LTCFs running resilience training sessions (29) or disseminating resources that aid the development of resilience such as the ‘*Resilience Resource*’ tool (22). It, however, must be noted that resilience is not entirely the responsibility of an individual, but resilience can be cultivated organisationally. A shift to focus more on improving organisational resilience is required within the care sector. This includes a much needed culture change in order to ensure that this sector is valued and recognised, making staff feel more appreciated. This will not only contribute to improved mental health amongst staff but it is likely to result in increased retention and staff cohesion (30). It is also recommended that this change extends to researchers wishing to undertake research in LTCFs; thus, the authors strongly recommend future researchers show appreciation for LTCFs and staff who take part in research – this could include financial incentives, donations, or gestures of gratitude (25).

### Strengths and limitations

This study has the advantage of incorporating a wide range of care staff’s voices, in terms of roles, within the context of the COVID-19 pandemic and the related restrictions regarding the impact on their mental and emotional wellbeing. In doing so, it has highlighted the importance of providing support to and encouraging resilience for those working in the care sector. It is the first study, to the authors’ knowledge, to explore UK LTCF staff’s emotional wellbeing in the context of COVID-19 restrictions and after restrictions were removed.

However, the study restricted recruitment to one region in England, and excluded LTCFs whose most recently received a CQC rating of ‘requiring improvement’. Future research would benefit from including LTCFs with lower CQC ratings as their experiences of the pandemic and its impact on longer-term wellbeing of staff may differ from LTCFs rated ‘good’ or ‘outstanding’. Moreover, the authors were unable to contact care staff who had left the sector during/after the pandemic. Individuals who left the care sector may also have contrasting experiences of working during the pandemic and the consequent impact on their emotional health. The study also did not collect any data on the mental health and wellbeing of participants prior to the pandemic, further limiting the conclusions that can be drawn.

## Conclusion

The impact of the COVID-19 pandemic has had severe and long-lasting consequences on LTCF staff’s emotional wellbeing. Urgent research is needed to co-develop interventions that are timely and appropriate to emotionally support LTCF staff, and to explore how LTCFs can be supported to promote organisational resilience for staff working in this sector. This is particularly important in terms of future pandemic planning as well as improving the emotional health of current care staff.

⍰

## Abbreviations

CHAP: Care Home Assistant Practitioner
CQC: Care Quality Commission
LTCF: Long-term care facility ⍰

## Ethics approval and consent to participate⍰

This study was performed in accordance with the Declaration of Helsinki. Research Ethics Committee approval was obtained from the Lancaster University Faculty of Health and Medicine Research Ethics Committee (reference: FHM-2023-3368-RECR-3). Written informed consent was collected from both the manager of the LTCF recruited to the study and the LTCF staff member participating in the interview for anonymised publication of the data collected.

⍰

## Consent for publication⍰

Not applicable. Participant identification numbers and removal of personal details ensure anonymity of those involved in the interviews.

## Data availability

Data is available on reasonable request from the authors.

⍰

## Funding⍰

The research was funded by the Dowager Countess Eleanor Peel Trust, as part of the Sir Robert Boyd Fellowship. The views expressed are those of the authors and not necessarily those of the Dowager Countess Eleanor Peel Trust. ⍰

⍰

## Authors’ contributions⍰

DCM conceived the study, developed the study protocol, applied for ethical approval, and prepared the manuscript. NC coordinated recruitment conducted the interviews and contributed to the overall writing of the paper. DCM and NC analysed the data. ⍰

## Acknowledgments

The authors would like to acknowledge and thank the Enabling Research in Care Homes network for supporting the recruitment for the study from which this data was collected. The authors would also like to thank the LTCFs and staff members who took part in this research for their valuable contribution.

